# Linear growth trajectories in the first two years of life predict attained linear growth and stunting at the age of five years: Results from the MAL-ED multi-country birth cohort study

**DOI:** 10.1101/2025.08.01.25332596

**Authors:** Md. Ashraful Alam, S. M. Tafsir Hasan, Amena Al Nishan, Mustafa Mahfuz, Margaret N Kosek, Aldo A. M. Lima, Bruna L. L. Maciel, Tahmeed Ahmed

**Affiliations:** Nutrition Research Division, International Centre for Diarrhoeal Disease Research, Bangladesh, (icddr,b), Dhaka 1212, Bangladesh; Poche Centre for Indigenous Health, The University of Queensland, Brisbane, QLD, Australia; Division of Infectious Diseases and International Health, School of Medicine, University of Virginia, Charlottesville, Virginia, USA; Universidade Federal do Ceará, Fortaleza, and Universidade Federal do Rio Grande do Norte, Natal, Brazil; Office of Executive Director, International Centre for Diarrhoeal Disease Research, Bangladesh, (icddr,b), Dhaka 1212, Bangladesh

**Author notes:** Correspondence S. M. Tafsir Hasan, MBBS, MS Associate Scientist, Nutrition Research Division, International Centre for Diarrhoeal Disease Research, Bangladesh (icddr,b).

**Keywords:** Latent class growth model, growth faltering, stunting, linear growth trajectory, early childhood

## Abstract

**Background:** Linear growth faltering and stunting are associated with childhood mortality, morbidity, and impaired growth and cognitive development.

**Objectives:** We sought to identify groups of children with different growth trajectories in their first two years of life and determine if there is an association between those early-life trajectories and attained linear growth and stunting at age five.

**Methods:** We used the MAL-ED birth cohort study’s dataset in this analysis. The latent class growth modeling (LCGM) technique was used to identify unique classes of children who followed similar trajectories in terms of length for age z-score (LAZ) during the age of 0 to 24 months. Mixed-effects linear and logistic regression models were used to investigate the association of the LCGM-derived trajectories with height for age z-score (HAZ) and stunting at age 60 months, respectively, considering the study site as the random effect.

**Results:** We detected five LAZ trajectories in 1471 children aged 0 to 2 years and designated them as follows: Class 1: severely attenuated linear growth (9%); class 2: moderately attenuated linear growth (25%); class 3: mildly attenuated linear growth (34%); class 4: stable linear growth (25%); class 5: improved linear growth (7%). In adjusted model, LAZ trajectories in the first 2 years of life were associated with HAZ and stunting at 5 years. Compared to the stable linear growth class, the improved linear growth class had a predicted 0.86 higher HAZ at age 5 years (95% CI: 0.67, 1.04), but the severely attenuated linear growth classes had lower HAZ at age 5 years (β = -2.10; 95% CI: -2.26, -1.95).

**Conclusions:** Linear growth trajectories during the first two years of life are crucial as they predict the attained linear growth and stunting at 5 years. Emphasis should be given to improving linear growth in early life through community interventions.

## INTRODUCTION

Linear growth faltering in children remains a major global health challenge, with a staggering 148.1 million stunted children under the age of 5 years in low and middle-income countries (LMICs) (1). Evidence suggests that early-life stunting is associated with an increased risk of childhood mortality (2), infectious diseases such as diarrhea, pneumonia, and measles (3, 4), impaired growth and cognitive development (5–8) as well as poor adult economic outcomes (9). Furthermore, recent studies have demonstrated an impact of impaired early-life linear growth on late childhood growth with infrequent reversal of stunting (10, 11).

While several nutritional interventions have shown substantial potential in addressing childhood stunting, achieving widespread success remains elusive (12). Moreover, growth during the first thousand days is influenced by several factors, including environment, infectious disease, and malnutrition, and is not homogeneous in pattern (13, 14). As different children follow different patterns of growth (15), characterizing childhood growth trajectories and implementing separate interventions for different groups has become essential. Novel strategies focused on specific growth pattern trajectories would place us in a better position to fight against childhood stunting.

Group trajectory models can designate a group of individuals with similar features, characteristics, and attributes into discrete, unambiguous subgroups (16–18) and have become expository in understanding patterns of child growth and development. Despite that some studies explored trends of linear growth in low-income settings (19, 20), comprehensive data is limited in LMICs regarding the presence of distinct group-based trajectories of linear growth among young children. It is also to be seen how much the early linear growth trajectories influence and set the course for linear growth attained later in childhood.

This study aimed to identify groups of children with distinct linear growth trajectories in the first two years of life in a birth cohort from seven different LMICs. This study also assessed the association of these early childhood linear growth trajectories with attained linear growth at age 60 months.

## SUBJECTS AND METHODS

### Ethics statement

The study was conducted in accordance with the guidelines of the Declaration of Helsinki. The study (Protocol ID: PR-2008-020) was approved by the Research Review Committee and the Ethical Review Committee (Institutional Review Board) of icddr,b. Written informed consent was obtained from the legal guardians of all participants.

### Study setting and participants

The MAL-ED study is a multi-country prospective community-based birth cohort study conducted at eight locations in LMICs (21). Data were collected between November 2009 and February 2017 in Fortaleza, Brazil (BRF); Dhaka, Bangladesh (BGD); Vellore, India (INV); Loreto, Peru (PEL); Bhaktapur, Nepal (NEB); Naushero Feroze, Pakistan (PKN); Haydom, Tanzania (TZH) and Venda, South Africa (SAV). We eliminated Pakistan from these analyses due to bias in length measures in a subgroup of subjects. Initially, each site planned to enroll and follow 200 children for the next 24 months. Eventually, through an amendment to the original protocol, the children were followed up to the age of 60 months. Eligible newborns were fewer than 17 days old, born singletons with a birth weight > 1500 g, free of significant illnesses, born to a mother who was at least 16 years old, and who planned to stay in the community for at least 6 months.

### Anthropometry, dietary intake, and socioeconomic status

From birth to 60 months of age, trained field staff measured children’s recumbent length (using a Seca 417 Infantometer) or standing height (using a Seca 213 portable stadiometer) with a precision of 0.1 cm monthly following standard procedures. We derived length/height-for-age z scores using the WHO growth standards (22). Stunting at age 60 months was defined as a height-for-age z score less than -2.

Trained field workers determined the dietary intake using quantitative 24-hour recalls at 60 months (23). Subsequently, the dietary intake data underwent conversion to quantify energy, macronutrient, and micronutrient intakes from non-breast milk foods, employing site-specific food composition databases developed within MAL-ED.

The MAL-ED study developed a composite socioeconomic status (SES) score that could be compared across study locations. The SES construct combines improved access to water and sanitation, maternal education, assets, and average monthly household income to provide a score (WAMI) that goes from 0 to 1 (24).

### Statistical Analysis

During the age of 0 to 24 months, the latent class growth modeling (LCGM) technique was used to identify unique classes or clusters of children who followed similar trajectories in terms of length for age z score (LAZ). LCGM is a semi-parametric finite mixture modeling technique that uses maximum likelihood to evaluate longitudinal data to identify distinct and meaningful groups of people that have analogous progression through time for a particular variable. The assumption that all individuals come from a single population is relaxed in LCGM, which allows for differences in growth parameters between unobserved subpopulations. However, it presumes that the slope and intercept are constant for all members of each different group (25, 26).

For LAZ, we created distinct trajectory models. We had a total of 25 measurement time points (to provide acceptable parameter estimates of trajectories, LCGM required at least three measurement time-points for each case) (27). This analysis included only children with 0- and 24-month outcome data.

We developed and compared different models with 1 to 5 trajectories for the outcome. LAZ was modeled using a censored normal distribution technique. We fit models of increasing complexity starting with a single trajectory and chose the model that best suited the data to find the optimal number of trajectories. Linear, quadratic, cubic, and quartic functions of time (age in months) were explored to find the unique trajectories of the outcomes. Models that produced trajectories with insufficient cluster size (less than 5% of the study population) were not considered.

Based on Bayesian information criteria (BIC), log Bayes factor, the statistical significance of cubic terms, whether 95 percent confidence intervals of trajectories overlapped, and the percentage of the population in each trajectory group, we selected the final models with the optimal number and shape of trajectories. The BIC with the smallest absolute value showed the best fit (28). After selecting the final model, we estimated the posterior probabilities for each individual belonging to each of the trajectory groups. Children were assigned to a trajectory group using the maximum probability assignment criterion (29).

To determine the final models’ goodness of fit, we looked at whether the average posterior probability of assignment for each subgroup was greater than 0.8, whether the odds of correct classification were greater than 5, and whether the estimated/modeled group probabilities were in good agreement with the proportions of group assignments) (27). Following *the GRoLTS- Checklist: Guidelines for Reporting on Latent Trajectory Studies*, we reported the results of the LCGM (30).

A linear mixed-effects model was used to investigate the relationship between HAZ at the age of 60 months and the trajectories found by LCGM, including a random intercept for the research site. We included the site as a random effect to account for clustering by site. This method enables for accurate estimation of the outcome variable’s variation both within and between clusters (31).

Additionally, a mixed-effects logistic regression model was applied to assess the relationship between stunting at 60 months and the LCGM trajectories, also including a random intercept for the research site. Due to the small number of stunted children, classes 3, 4, and 5 of the LAZ trajectory groups were combined for the analysis. To construct these models, we used data collected at the age of 60 months, including the sum of protein in grams for the day of food recall, the sum of fat in grams for the day of food recall, the sum of carbohydrates in grams for the day of food recall, socioeconomic status (WAMI score), and child sex. The strength of association was expressed as mean difference (β) and odds ratio (OR) with 95% confidence interval (95% CI) for the linear and logit models, respectively.

The “traj plugin” in Stata was used to conduct the statistical analyses relating to LCGM (32), a Stata equivalent of the widely used “proc traj” in SAS (28). The R packages “lcmm” and “ggplot2” were used to plot the outputs of LCGM models. Stata 15.0 was used for all other statistical analyses.

## RESULTS

In the MAL-ED study, we enrolled 1,868 children from seven countries (Bangladesh, India, Nepal, Brazil, Peru, South Africa, and Tanzania). Of these, 1469 children had length data at birth and 24 months, and 1047 children had complete data on all other variables (**Figure 1**).

**Figure 1.**
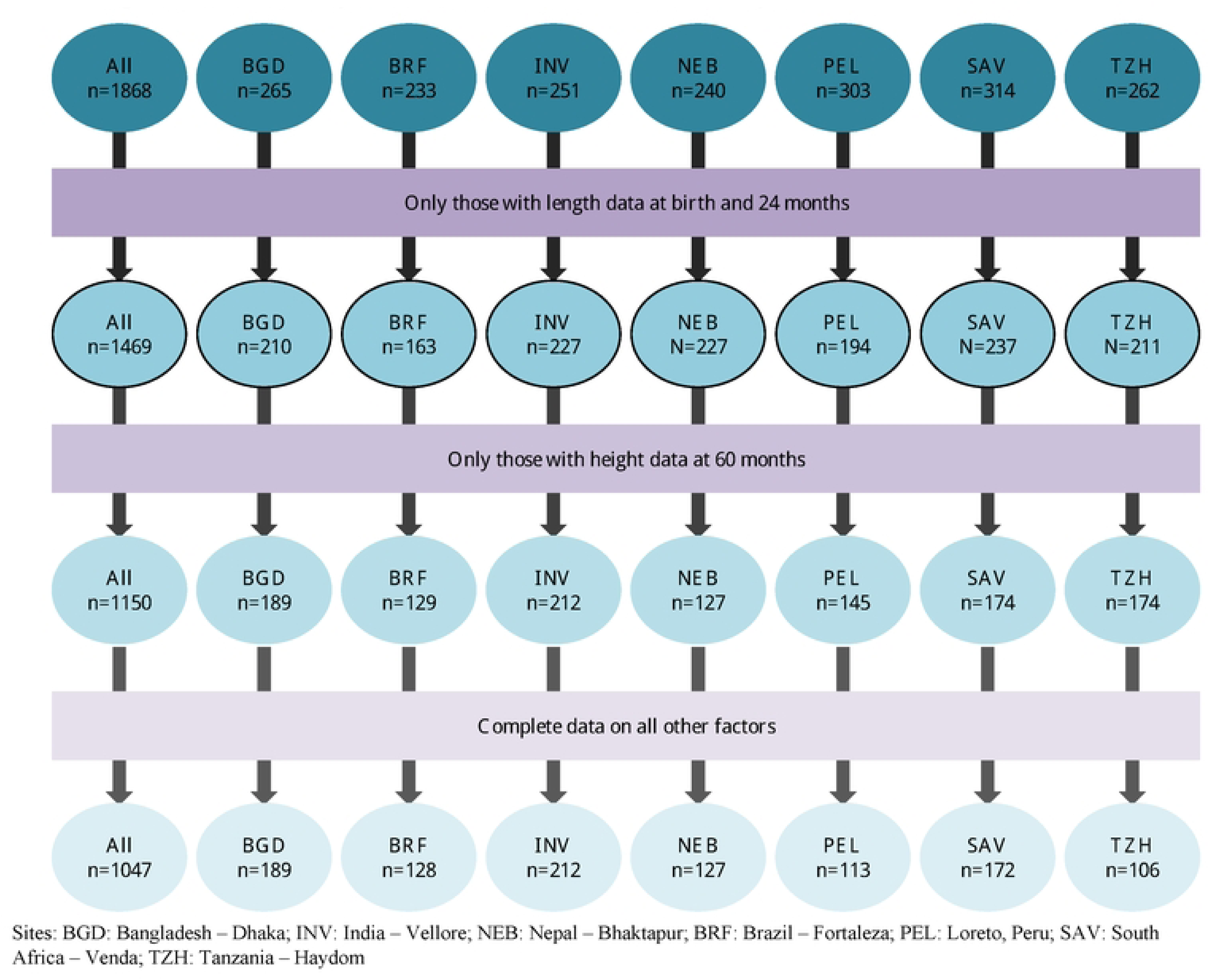
MAL-ED cohort profile of children who were retained in the analysis. [Sites: BGD: Bangladesh – Dhaka; INV: India – Vellore; NEB: Nepal – Bhaktapur; BRF: Brazil – Fortaleza; PEL: Loreto, Peru; SAV: South Africa – Venda; TZH: Tanzania – Haydom]

Of the children, 49% were girls. The average length-for-age z score at birth ranged from -1.02 in INV and TZH to -0.71 in SAV. Overall, median protein consumption was 37.7 g/d with the lowest median protein intake in NEB (30.4 g/d) and the highest median protein intake in BRF (54.0 g/d) at 60 months of age. The seven locations had different median fat intakes, ranging from 21.3 g/d in TZH to 45.4 g/d in BRF. The carbohydrate intakes in the seven areas varied, ranging from 171 g/d in NEB to 318 g/d in TZH (**Table 1**).

**Table 1:**
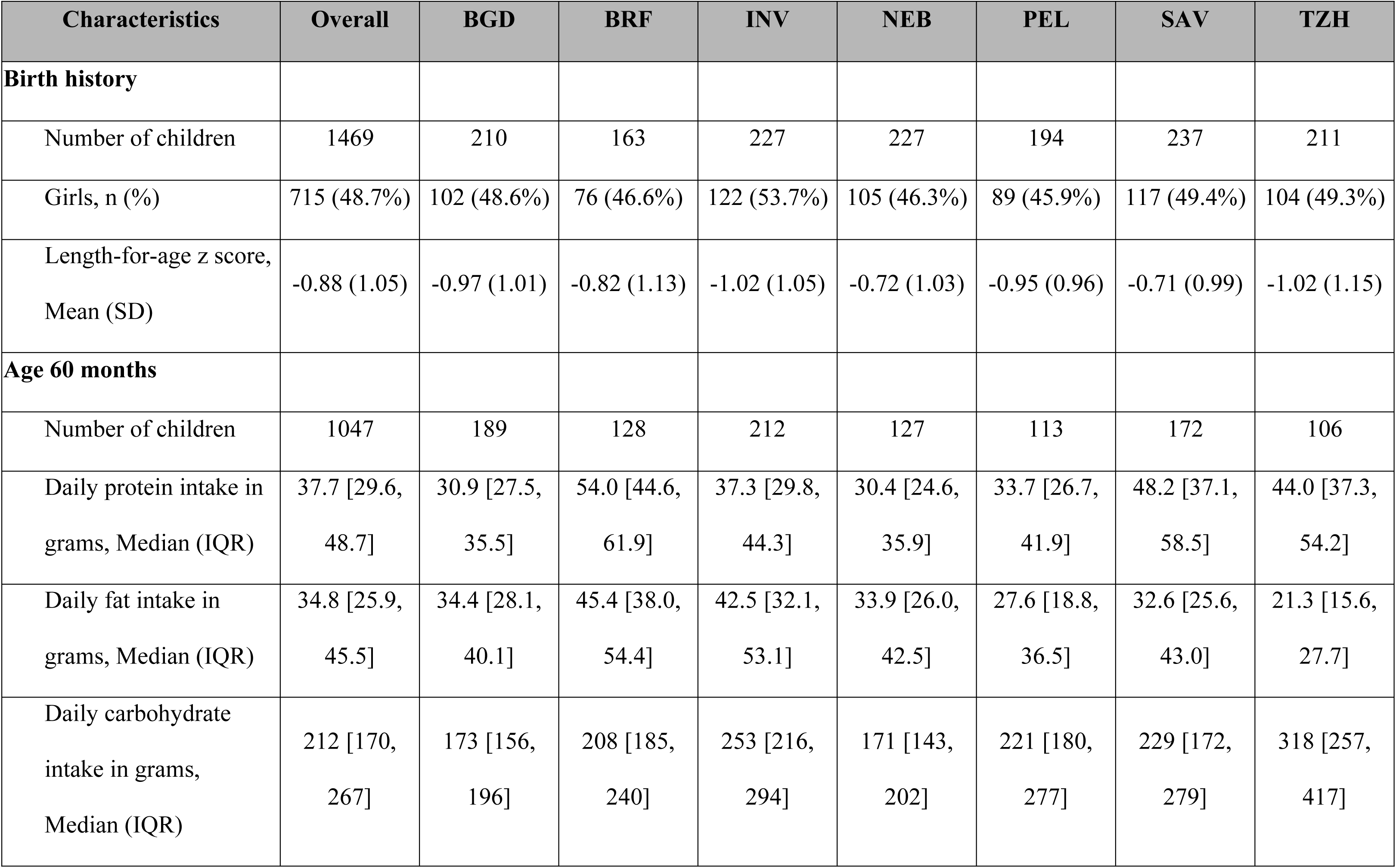
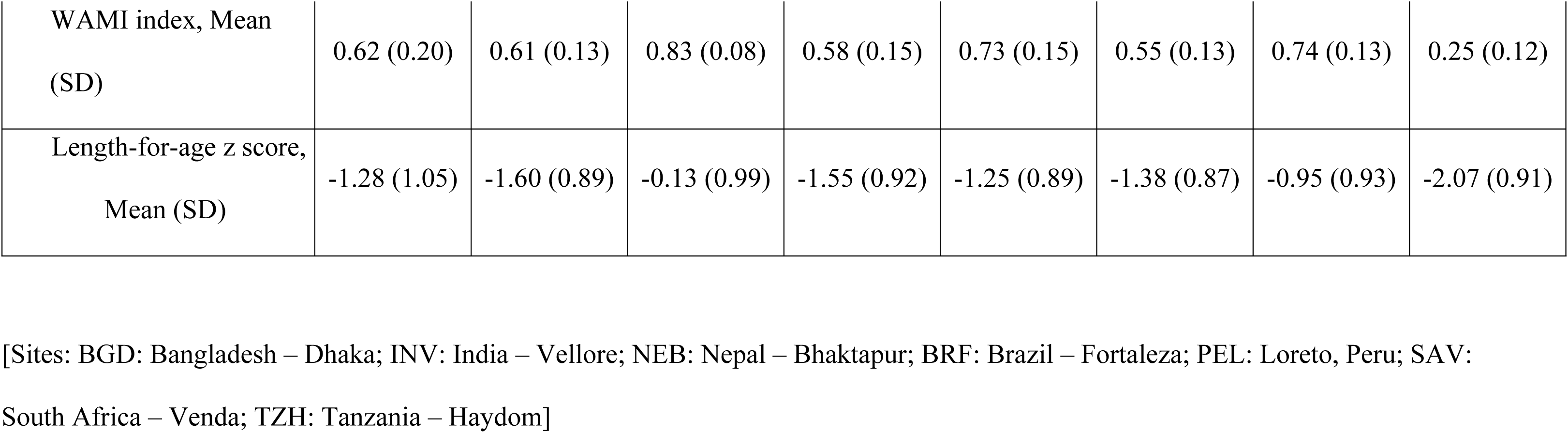
Study population characteristics.

| Characteristics | Overall | BGD | BRF | INV | NEB | PEL | SAV | TZH |
| --- | --- | --- | --- | --- | --- | --- | --- | --- |
| **Birth history** |  |  |  |  |  |  |  |  |
| Number of children | 1469 | 210 | 163 | 227 | 227 | 194 | 237 | 211 |
| Girls, n (%) | 715 (48.7%) | 102 (48.6%) | 76 (46.6%) | 122 (53.7%) | 105 (46.3%) | 89 (45.9%) | 117 (49.4%) | 104 (49.3%) |
| Length-for-age z score,<br>Mean (SD) | -0.88 (1.05) | -0.97 (1.01) | -0.82 (1.13) | -1.02 (1.05) | -0.72 (1.03) | -0.95 (0.96) | -0.71 (0.99) | -1.02 (1.15) |
| **Age 60 months** |  |  |  |  |  |  |  |  |
| Number of children | 1047 | 189 | 128 | 212 | 127 | 113 | 172 | 106 |
| Daily protein intake in<br>grams, Median (IQR) | 37.7 [29.6,<br>48.7] | 30.9 [27.5,<br>35.5] | 54.0 [44.6,<br>61.9] | 37.3 [29.8,<br>44.3] | 30.4 [24.6,<br>35.9] | 33.7 [26.7,<br>41.9] | 48.2 [37.1,<br>58.5] | 44.0 [37.3,<br>54.2] |
| Daily fat intake in<br>grams, Median (IQR) | 34.8 [25.9,<br>45.5] | 34.4 [28.1,<br>40.1] | 45.4 [38.0,<br>54.4] | 42.5 [32.1,<br>53.1] | 33.9 [26.0,<br>42.5] | 27.6 [18.8,<br>36.5] | 32.6 [25.6,<br>43.0] | 21.3 [15.6,<br>27.7] |
| Daily carbohydrate<br>intake in grams,<br>Median (IQR) | 212 [170,<br>267] | 173 [156,<br>196] | 208 [185,<br>240] | 253 [216,<br>294] | 171 [143,<br>202] | 221 [180,<br>277] | 229 [172,<br>279] | 318 [257,<br>417] |
| WAMI index, Mean<br>(SD) | 0.62 (0.20) | 0.61 (0.13) | 0.83 (0.08) | 0.58 (0.15) | 0.73 (0.15) | 0.55 (0.13) | 0.74 (0.13) | 0.25 (0.12) |
| Length-for-age z score,<br>Mean (SD) | -1.28 (1.05) | -1.60 (0.89) | -0.13 (0.99) | -1.55 (0.92) | -1.25 (0.89) | -1.38 (0.87) | -0.95 (0.93) | -2.07 (0.91) |

[Sites: BGD: Bangladesh – Dhaka; INV: India – Vellore; NEB: Nepal – Bhaktapur; BRF: Brazil – Fortaleza; PEL: Loreto, Peru; SAV:
South Africa – Venda; TZH: Tanzania – Haydom]

### Results of LCGM

The difference in the population distribution of the progression of LAZ was best defined by a five-class model with a cubic component for the first three trajectories and quartic components for the other two trajectories. The detected LAZ trajectory class 1, termed “severely attenuated linear growth,” which represented 9% of the children in our sample, was born with a mean LAZ close to -2.15 and had a notable drop in LAZ by the time they were 24 months old (mean LAZ=-2.73, -3.25, -3.60, -3.69 at 6m, 12m, 18 and 24m, respectively). Class 2, named “moderately attenuated linear growth,” accounted for 25% of the participants, and these children’s mean LAZ was -1.37 at birth and decreasing up to 24 months (LAZ=-1.76, -2.19, - 2.50, -2.56 at 6m, 12m, 18 and 24m, respectively). Trajectory class 3, termed “mildly attenuated linear growth,” accounted for 34% of the participants, this group on average was close to -0.84 at birth and slightly decreased until 24 months of age. Trajectory class 4, termed “stable linear growth,” made up 25% of the participants, and their LAZ slightly increased during the first five months before falling over the next 24 months. LAZ trajectory class 5, named “improved linear growth,” which accounted for 7% of the children, was born with a mean LAZ near -0.05, grew significantly until 12 months of age, then decreased slightly until 24 months (LAZ=0.82, 0.91, 0.87, 0.57 at 6m, 12m, 18 and 24m, respectively) (**Figure 2**) (**Table 2**).

**Figure 2.**
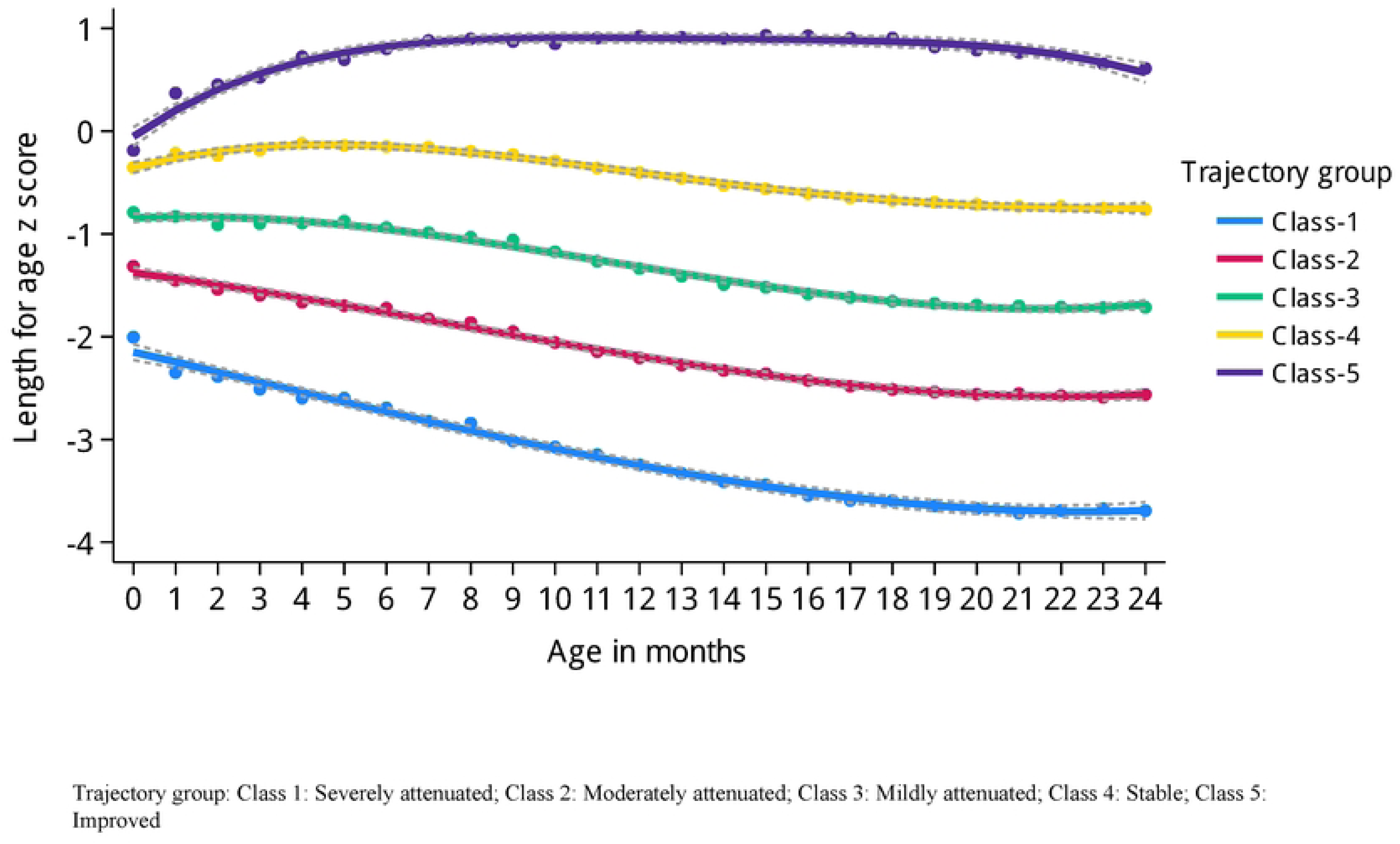
LCGM derived latent trajectories with 95% confidence interval of length-for- age z-score during age 0 to 24 months. [Trajectory group: Class 1: Severely attenuated; Class 2: Moderately attenuated; Class 3: Mildly attenuated; Class 4: Stable; Class 5: Improved]

**Table 2.**
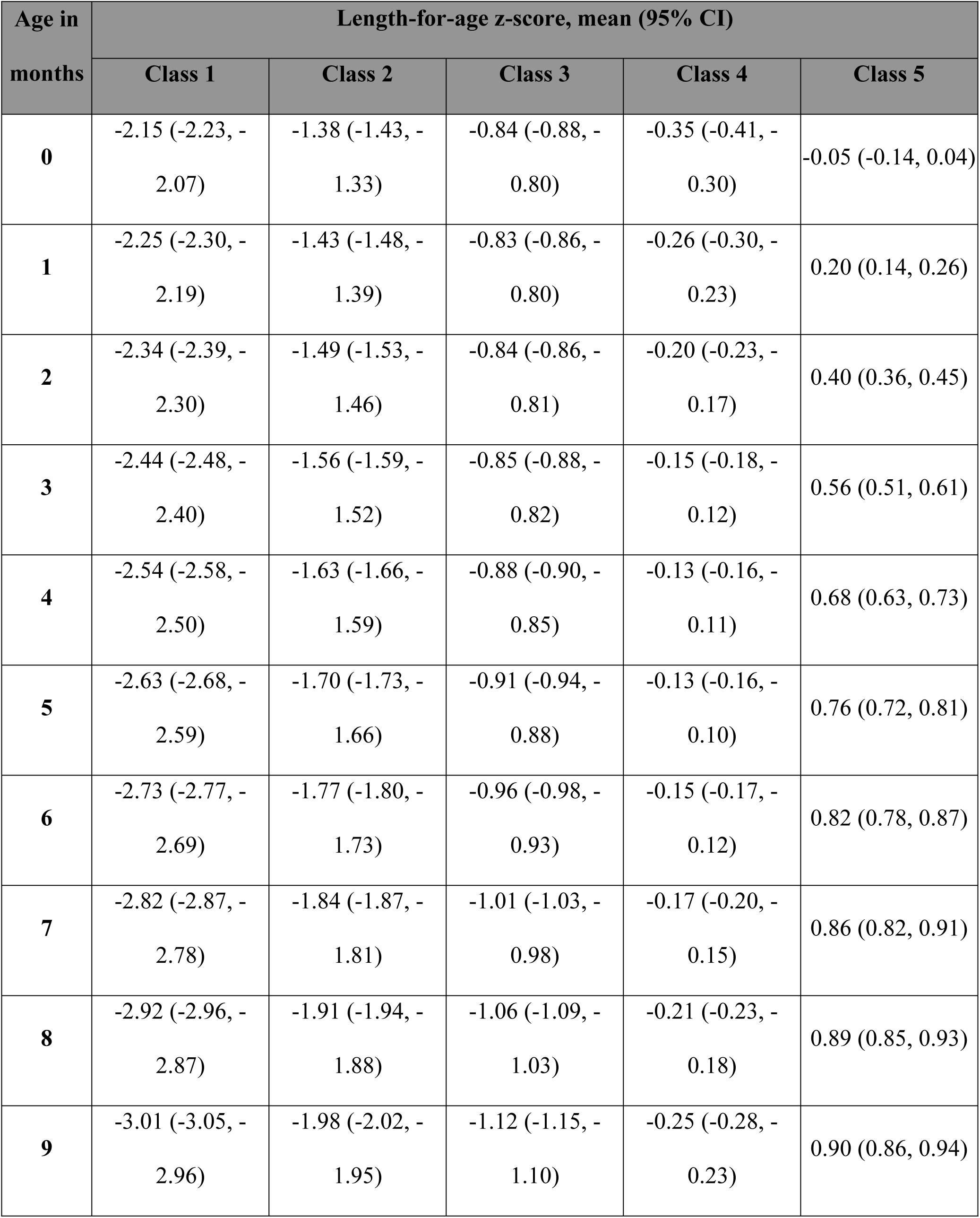
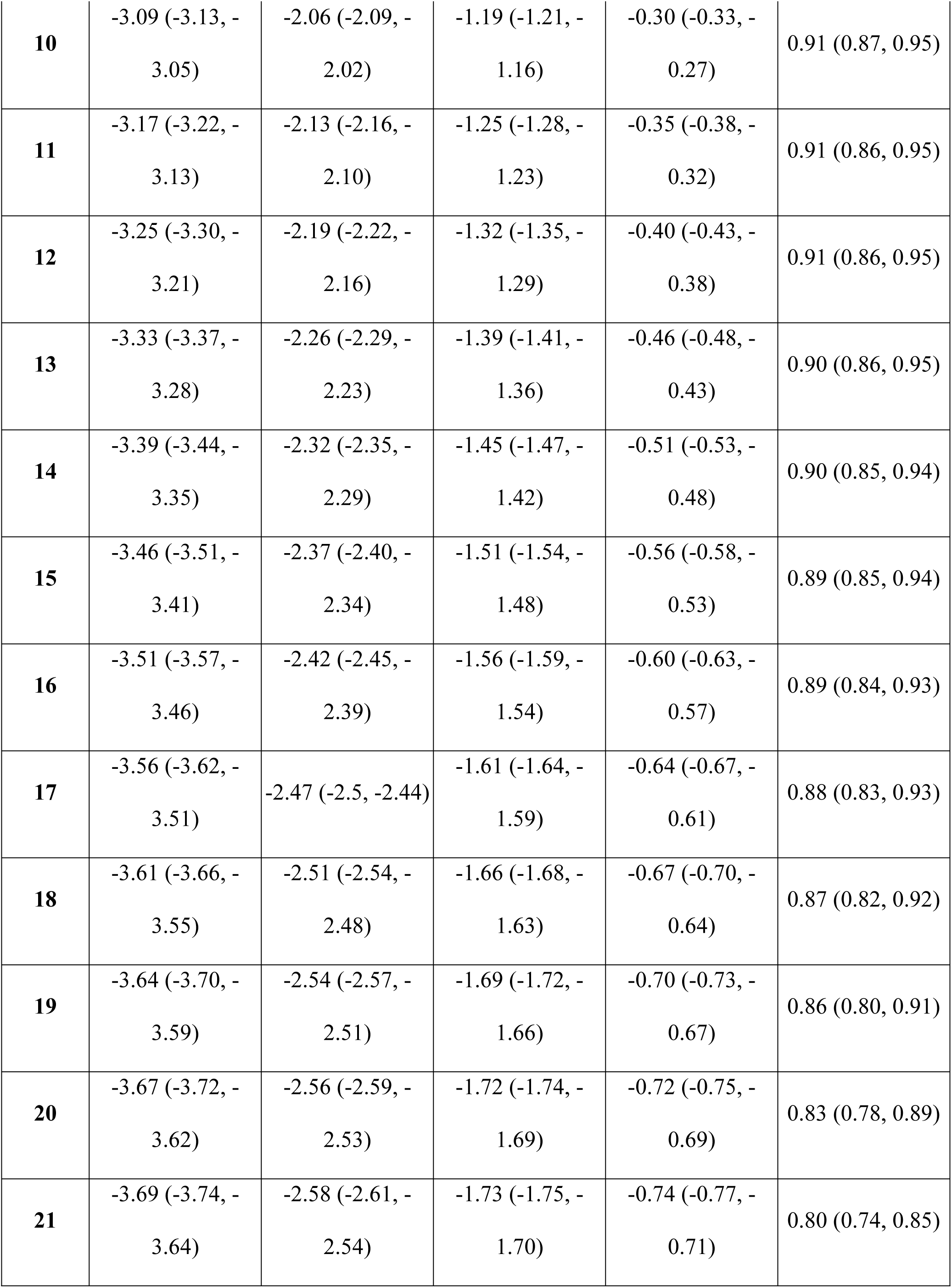
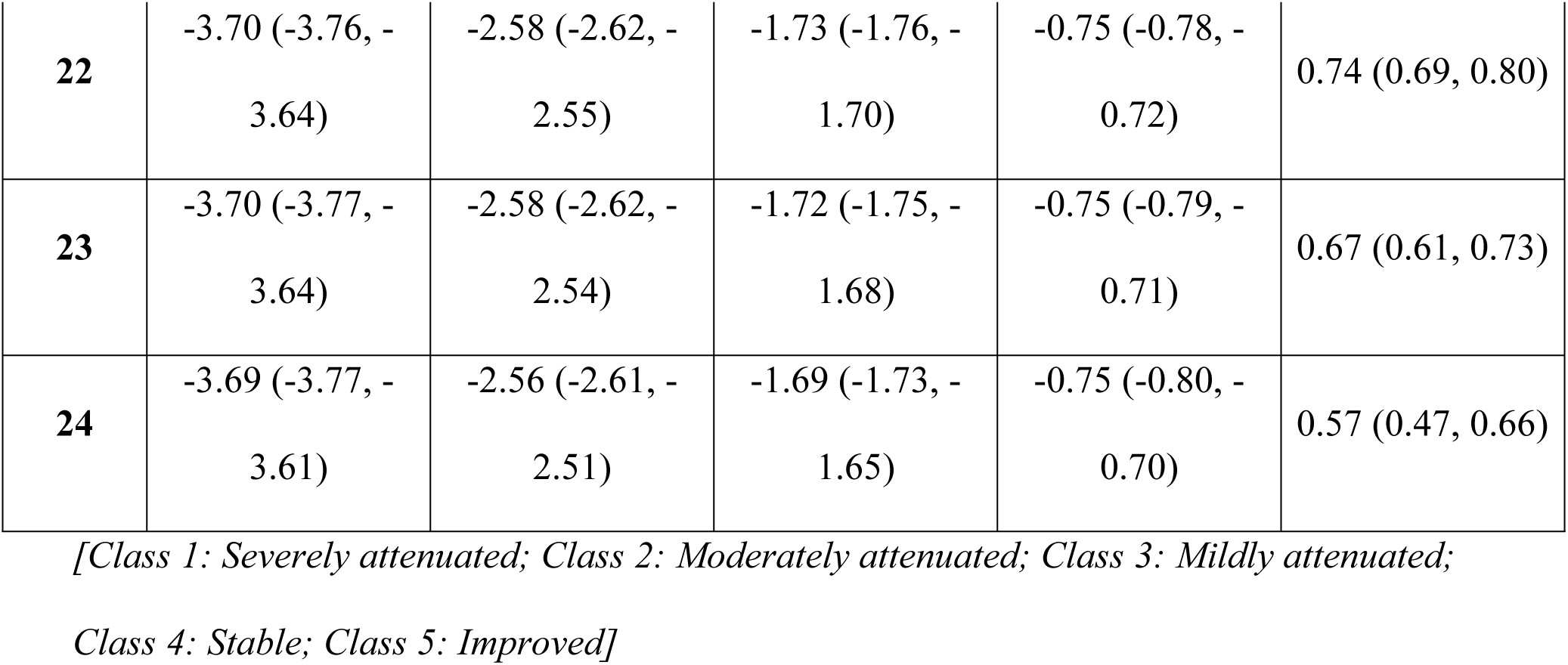
Trajectory-wise estimated mean with 95% confidence interval for length-for- age z-score at different ages derived using LCGM.

| Age in<br>months | Length-for-age z-score, mean (95% CI) |  |  |  |  |
| --- | --- | --- | --- | --- | --- |
|  | Class 1 | Class 2 | Class 3 | Class 4 | Class 5 |
| **0** | -2.15 (-2.23, -<br>2.07) | -1.38 (-1.43, -<br>1.33) | -0.84 (-0.88, -<br>0.80) | -0.35 (-0.41, -<br>0.30) | -0.05 (-0.14, 0.04) |
| **1** | -2.25 (-2.30, -<br>2.19) | -1.43 (-1.48, -<br>1.39) | -0.83 (-0.86, -<br>0.80) | -0.26 (-0.30, -<br>0.23) | 0.20 (0.14, 0.26) |
| **2** | -2.34 (-2.39, -<br>2.30) | -1.49 (-1.53, -<br>1.46) | -0.84 (-0.86, -<br>0.81) | -0.20 (-0.23, -<br>0.17) | 0.40 (0.36, 0.45) |
| **3** | -2.44 (-2.48, -<br>2.40) | -1.56 (-1.59, -<br>1.52) | -0.85 (-0.88, -<br>0.82) | -0.15 (-0.18, -<br>0.12) | 0.56 (0.51, 0.61) |
| **4** | -2.54 (-2.58, -<br>2.50) | -1.63 (-1.66, -<br>1.59) | -0.88 (-0.90, -<br>0.85) | -0.13 (-0.16, -<br>0.11) | 0.68 (0.63, 0.73) |
| **5** | -2.63 (-2.68, -<br>2.59) | -1.70 (-1.73, -<br>1.66) | -0.91 (-0.94, -<br>0.88) | -0.13 (-0.16, -<br>0.10) | 0.76 (0.72, 0.81) |
| **6** | -2.73 (-2.77, -<br>2.69) | -1.77 (-1.80, -<br>1.73) | -0.96 (-0.98, -<br>0.93) | -0.15 (-0.17, -<br>0.12) | 0.82 (0.78, 0.87) |
| **7** | -2.82 (-2.87, -<br>2.78) | -1.84 (-1.87, -<br>1.81) | -1.01 (-1.03, -<br>0.98) | -0.17 (-0.20, -<br>0.15) | 0.86 (0.82, 0.91) |
| **8** | -2.92 (-2.96, -<br>2.87) | -1.91 (-1.94, -<br>1.88) | -1.06 (-1.09, -<br>1.03) | -0.21 (-0.23, -<br>0.18) | 0.89 (0.85, 0.93) |
| **9** | -3.01 (-3.05, -<br>2.96) | -1.98 (-2.02, -<br>1.95) | -1.12 (-1.15, -<br>1.10) | -0.25 (-0.28, -<br>0.23) | 0.90 (0.86, 0.94) |
| **10** | -3.09 (-3.13, -<br>3.05) | -2.06 (-2.09, -<br>2.02) | -1.19 (-1.21, -<br>1.16) | -0.30 (-0.33, -<br>0.27) | 0.91 (0.87, 0.95) |
| **11** | -3.17 (-3.22, -<br>3.13) | -2.13 (-2.16, -<br>2.10) | -1.25 (-1.28, -<br>1.23) | -0.35 (-0.38, -<br>0.32) | 0.91 (0.86, 0.95) |
| **12** | -3.25 (-3.30, -<br>3.21) | -2.19 (-2.22, -<br>2.16) | -1.32 (-1.35, -<br>1.29) | -0.40 (-0.43, -<br>0.38) | 0.91 (0.86, 0.95) |
| **13** | -3.33 (-3.37, -<br>3.28) | -2.26 (-2.29, -<br>2.23) | -1.39 (-1.41, -<br>1.36) | -0.46 (-0.48, -<br>0.43) | 0.90 (0.86, 0.95) |
| **14** | -3.39 (-3.44, -<br>3.35) | -2.32 (-2.35, -<br>2.29) | -1.45 (-1.47, -<br>1.42) | -0.51 (-0.53, -<br>0.48) | 0.90 (0.85, 0.94) |
| **15** | -3.46 (-3.51, -<br>3.41) | -2.37 (-2.40, -<br>2.34) | -1.51 (-1.54, -<br>1.48) | -0.56 (-0.58, -<br>0.53) | 0.89 (0.85, 0.94) |
| **16** | -3.51 (-3.57, -<br>3.46) | -2.42 (-2.45, -<br>2.39) | -1.56 (-1.59, -<br>1.54) | -0.60 (-0.63, -<br>0.57) | 0.89 (0.84, 0.93) |
| **17** | -3.56 (-3.62, -<br>3.51) | -2.47 (-2.5, -2.44) | -1.61 (-1.64, -<br>1.59) | -0.64 (-0.67, -<br>0.61) | 0.88 (0.83, 0.93) |
| **18** | -3.61 (-3.66, -<br>3.55) | -2.51 (-2.54, -<br>2.48) | -1.66 (-1.68, -<br>1.63) | -0.67 (-0.70, -<br>0.64) | 0.87 (0.82, 0.92) |
| **19** | -3.64 (-3.70, -<br>3.59) | -2.54 (-2.57, -<br>2.51) | -1.69 (-1.72, -<br>1.66) | -0.70 (-0.73, -<br>0.67) | 0.86 (0.80, 0.91) |
| **20** | -3.67 (-3.72, -<br>3.62) | -2.56 (-2.59, -<br>2.53) | -1.72 (-1.74, -<br>1.69) | -0.72 (-0.75, -<br>0.69) | 0.83 (0.78, 0.89) |
| **21** | -3.69 (-3.74, -<br>3.64) | -2.58 (-2.61, -<br>2.54) | -1.73 (-1.75, -<br>1.70) | -0.74 (-0.77, -<br>0.71) | 0.80 (0.74, 0.85) |
| **22** | -3.70 (-3.76, -<br>3.64) | -2.58 (-2.62, -<br>2.55) | -1.73 (-1.76, -<br>1.70) | -0.75 (-0.78, -<br>0.72) | 0.74 (0.69, 0.80) |
| **23** | -3.70 (-3.77, -<br>3.64) | -2.58 (-2.62, -<br>2.54) | -1.72 (-1.75, -<br>1.68) | -0.75 (-0.79, -<br>0.71) | 0.67 (0.61, 0.73) |
| **24** | -3.69 (-3.77, -<br>3.61) | -2.56 (-2.61, -<br>2.51) | -1.69 (-1.73, -<br>1.65) | -0.75 (-0.80, -<br>0.70) | 0.57 (0.47, 0.66) |

*[Class 1: Severely attenuated; Class 2: Moderately attenuated; Class 3: Mildly attenuated;*
*Class 4: Stable; Class 5: Improved]*

For all the subgroups of LAZ, the average posterior probability of group membership was greater than 0.90, odds of correct classification were greater than 50, and modeled group probabilities were in good agreement with the proportions of group assignment following the maximum-probability assignment rule (**Figure 3**) (**Table 3**).

**Figure 3:**
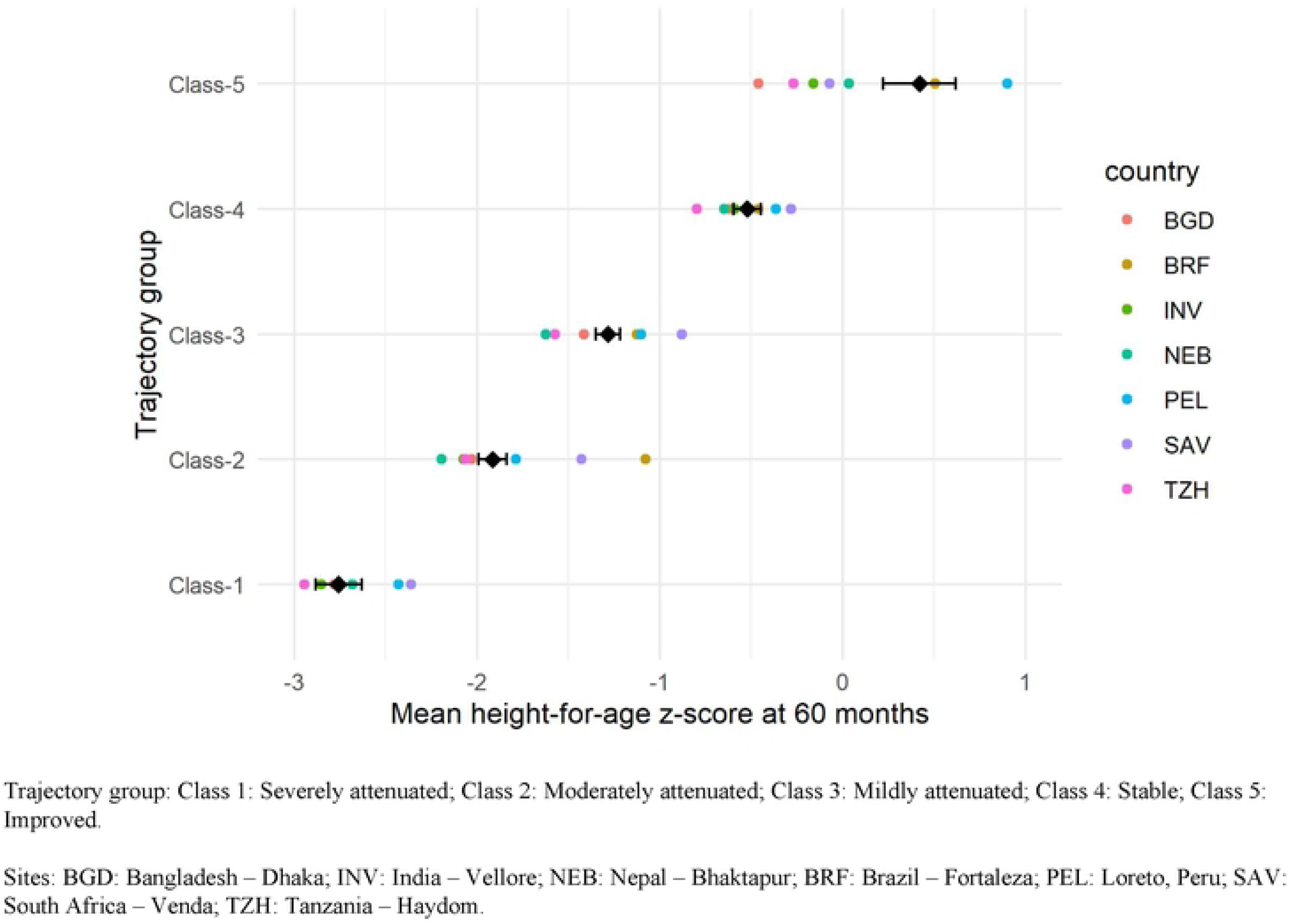
M**e**an **height-for-age z-score at 60 months by trajectory group over seven country sites.** [Trajectory group: Class 1: Severely attenuated; Class 2: Moderately attenuated; Class 3: Mildly attenuated; Class 4: Stable; Class 5: Improved]

**Table 3.** Fit statistics of the final models derived using LCGM for LAZ.

| LAZ trajectory group | n | Average<br>posterior<br>probability | Odds of<br>correct<br>classification | Estimated<br>group<br>probability | Proportion<br>of group<br>assignment |
| --- | --- | --- | --- | --- | --- |
| Class 1: Severely attenuated | 132 | 0.98 | 523.93 | 0.09 | 0.09 |
| Class 2: Moderately attenuated | 373 | 0.98 | 139.93 | 0.26 | 0.25 |
| Class 3: Mildly attenuated | 495 | 0.97 | 71.38 | 0.33 | 0.34 |
| Class 4: Stable | 363 | 0.98 | 176.25 | 0.25 | 0.25 |
| Class 5: Improved | 106 | 0.99 | 2706.68 | 0.07 | 0.07 |

### Effect of LAZ trajectories on HAZ at 5 years

LAZ trajectories in the first two years of life were associated with HAZ at the age of five (**Table 4**).

**Table 4:** Effect of the first two-year LAZ trajectories on height-for-age z-score at age 60 months (n= 1047)

| LAZ trajectory group | Height-for-age z-score |  |  |  |
| --- | --- | --- | --- | --- |
|  | Unadjusted |  | Adjusted |  |
| | $\beta$ (95% CI) | p-value | $\beta$ (95% CI) | p-value |
| Class 1: Severely attenuated | -2.14 (-2.30, -1.98) | <0.001 | -2.10 (-2.26, -1.94) | <0.001 |
| Class 2: Moderately attenuated | -1.37 (-1.49, -1.26) | <0.001 | -1.34 (-1.45, -1.23) | <0.001 |
| Class 3: Mildly attenuated | -0.73 (-0.84, -0.62) | <0.001 | -0.71 (-0.81, -0.60) | <0.001 |
| Class 4: Stable | Reference |  | Reference |  |
| Class 5: Improved | 0.86 (0.67, 1.05) | <0.001 | 0.86 (0.67, 1.04) | <0.001 |

*Adjusted for child sex, daily average fat, protein and carbohydrate intake and well as socio-* *economic status (WAMI)*

After adjusting for potential confounders, the improved linear growth class had a predicted 0.86 higher HAZ at age five years (95% CI: 0.67, 1.04), but the severely, moderately, and mildly attenuated linear growth classes had lower HAZ at age five years (severely: β = -2.10; 95% CI: -2.26, -1.95; moderately: β = -1.34; 95% CI: -1.45, -1.22; mildly: β = -0.70; 95% CI: -0.81, -0.59) when compared to the stable linear growth class.

### Effect of LAZ trajectories on stunting at 5 years

LAZ trajectories in the first two years of life were strongly associated with stunting at the age of five (**Table 5**).

**Table 5:** Effect of the first two-year LAZ trajectories on stunting at age 60 months (n= 1047)

| LAZ trajectory group | Stunting |  |  |  |
| --- | --- | --- | --- | --- |
|  | Unadjusted |  | Adjusted |  |
|  | OR (95% CI) | p-value | OR (95% CI) | p-value |
| Class 1: Severely attenuated | 142.08 (67.24, 300.20) | <0.001 | 132.78 (61.58, 286.52) | <0.001 |
| Class 2: Moderately attenuated | 16.95 (10.91, 26.35) | <0.001 | 16.00 (10.28, 24.91) | <0.001 |
| Class 3: Mildly attenuated/Class 4: Stable/ Class 5: Improved | Reference |  | Reference |  |

*Adjusted for child sex, daily average fat, protein and carbohydrate intake and well as socioeconomic status (WAMI)*

After adjusting for potential confounders, the severely attenuated linear growth class (OR 132.78; 95% CI: 61.58, 286.52) and moderately attenuated linear growth class (OR 16.00; 95% CI: 10.28, 24.91) had higher odds of developing stunting at the age of five years compared to other three classes.

## DISCUSSION

To the best of our knowledge, the present study is the first to identify discrete linear growth trajectories in the first two years of life, along with their association with the attained linear growth in later phases of childhood in a multi-country birth cohort from low-income settings. We observed the presence of five distinct linear growth trajectories in children from birth to 24 months using latent class growth modeling. Namirembe et al. demonstrated four distinct linear growth trajectories from birth to 12 months of age among children in rural Uganda (33). Another study that longitudinally followed a birth cohort of HIV-unexposed children in Zimbabwe identified four distinct linear growth trajectories from birth through 2 years of age by clustering infants with similar growth patterns (34).

Our results suggest these growth trajectories effectively describe the linear growth patterns, similarities, and heterogeneities of the trajectories and the influence of these trajectories on attained linear growth later in childhood. The sustained and discrete pattern of these growth trajectories from birth up to 24 months and their association with attained linear growth later at 60 months implicates the importance of early life monitoring of growth faltering to prevent stunting. We found that the “severely attenuated linear growth” group had the lowest LAZ at birth, falling in the stunted category (mean LAZ -2.15), and through the next 24 months, the LAZ continued to decline, reaching the severely stunted category (mean LAZ -3.69). In contrast, the “improved linear growth” group started as non-stunted (mean LAZ -0.05 at birth) and further improved thereafter in terms of LAZ with a slight tapering in the end (mean LAZ 0.57 at 24 months).

However, both these growth trajectory classes represent relatively small percentages of our study population (9% and 7%, respectively), and the highest number of children (34%) falls into the trajectory of “mildly attenuated linear growth”. Although this group started as non-stunted (mean LAZ -0.84 at birth), their mean LAZ gradually declined and ended up being in the at-risk-of-stunting category at the end of 24 months (mean LAZ -1.69). The remaining two trajectories have the same percentage of children (25%). The trajectory “moderately attenuated linear growth”, started in the at-risk-of-stunting category (mean LAZ 1.38 at birth), which further declined throughout the 24 months and ended up being in the stunted category at the end of 24 months (mean LAZ -2.56). The children with the trajectory “stable linear growth,” maintained a somewhat stable linear growth and remained non-stunted throughout the 24 months.

Our study revealed that, amongst the five trajectory classes, two classes that were at the bottom in terms of mean LAZ at birth, namely the “severely attenuated linear growth” and “moderately attenuated linear growth” developed stunting by 24 months. Our results showed a very high odds of developing stunting at the age of 5 years in the severely attenuated and moderately attenuated growth class trajectories compared to the other three classes. Complementing this finding, Krebs et al. stated that deficits in birth length could be the strongest predictors of attained LAZ and of stunting at the age of two years for the children from all four sites of their study (Democratic Republic of Congo, Guatemala, India, and Pakistan). The authors found that mild and moderate stunting at birth were associated with 0.5 to 1.0 lower adjusted mean difference in LAZ and a 40–60% increase in stunting risk at 2 years (35).

In addition to pointing out the effect of birth length on later phases of childhood, our study also hinted at a graded effect of the five trajectories on the attained linear growth at five years of age. We found that compared to the stable linear growth class, the mildly, moderately, and severely attenuated linear growth classes were associated with reduced HAZ at five years of age following a dose-response pattern. Also, the improved linear growth class was associated with increased HAZ in children at the age of five compared to the stable linear growth class. The underlying mechanism of these graded effects of early-life linear growth trajectories remains elusive. However, it can be somewhat explained by several studies that implicate the long-term and lifelong effects of early life growth faltering on the overall health outcome of an individual (36, 37). Furthermore, nutrition in the first 1000 days of life plays a pivotal role in shaping growth and development throughout one’s entire life (38, 39). Interestingly, our study demonstrated a disparity in daily consumption of carbohydrates, proteins, and fats among five-year-old children and the socioeconomic status (WAMI) across the seven sites.

All five trajectories detected in our study showed different patterns of linear growth, which stresses the importance of tracking and classifying growth patterns from birth and implementing effective group-specific targeted interventions to prevent growth faltering in children. A deep understanding of distinct early-life growth patterns is critical to promoting healthy growth in children because interventions based on population averages may not work for all. The timing of growth faltering, recovery patterns, and other determinants shaping the growth of children should be taken into consideration while developing strategies and tools to improve child health and growth. We recommend developing and implementing targeted novel strategies and health policies considering the children’s specific linear growth trajectories along with nutritional status, WASH environment, and socioeconomic status, which is also in line with advocacies from other policy recommendations (40, 41).

Although this study characterized the pattern of linear growth of children in early childhood and showed its association with attained linear growth in a later stage of childhood, it did not shed light on the factors determining the trajectories. An important limitation of this study is the lack of sufficient information on maternal stature, malnutrition, gestational weight gain, infections, and placental stress or insufficiency. Future research may examine the putative causes of linear growth faltering and stunting to explain the underlying factors and causality.

## CONCLUSION

This study demonstrated five distinct linear growth trajectories in the first two years of life among children from seven LMICs, ranging from severely attenuated to improved linear growth. The linear growth trajectories predicted attained linear growth at five years of age. Poor birth length and linear growth patterns in early childhood may lead to stunting in later stages of childhood. Public health measures should focus on identifying different classes of children with variable vulnerabilities and taking initiatives for group-based targeted interventions to eliminate linear growth faltering and stunting.

## Data Availability

All aggregated data related to this study are provided in the paper. To protect the identification of participants derived from the composite of key study variables, some restrictions do apply to the primary data. These data can be made available from the Ethics Committees (RRC and ERC) of icddr,b for researchers who meet the criteria for access to confidential data. Readers may contact the Senior Manager of Research Administration at icddr,b (Shiblee Sayeed) for data policies and queries.

## Funding

This work is based on the data from the “Etiology, Risk Factors, and Interactions of Enteric Infections and Malnutrition and the Consequences for Child Health and Development” (MAL-ED) project, which was funded by the Bill & Melinda Gates Foundation, the Foundation for the National Institutes of Health, and the National Institutes of Health’s Fogarty International Center. However, the authors received no funding for this work.

## Disclaimer

Notably, the funding bodies were not involved in the design of the study, data collection and analysis, decisions regarding publication, or the preparation of the manuscript.

## Competing interests

The authors declare that they have no competing interests.

## CONTRIBUTORS/ACKNOWLEDGEMENTS

MAA and SMTH designed research; MAA analyzed data; MAA, SMTH, and AAN wrote the manuscript; MM, MNK, AAML, BLLM, and TA reviewed and provided essential additional insights to the manuscript. MAA and SMTH had primary responsibility for final content. All authors read and approved the final manuscript.

## SUPPORTING INFORMATION

A STROBE Statement—Checklist of items is provided in Supplemental File 1.

## Notes

### Competing Interest Statement

The authors have declared no competing interest.

### Clinical Trial

NA

### Funding Statement

The author(s) received no specific funding for this work.

